## Supplemental Tables 1-7 for "Contributions of neighborhood social environment and air pollution exposure to Black-White disparities in epigenetic aging"

**S1 Table. Racial disparity in DNAm aging with weights or ancestry-informative principal components.**

| Characteristic | White, N = 2,438[1] | Black, N = 522[1] | Difference[2] | p-value[2] | Effect Size[3] |
|---|---|---|---|---|---|
| GrimAge raw residual | -0.34 (4.68) | 1.09 (4.86) | -1.4 | <0.001 | 0.299 |
| GrimAge weighted residual | -0.12 (4.68) | 1.33 (4.86) | -1.4 | <0.001 | 0.304 |
| GrimAge residual with ancestry-informative PCs | -0.34 (4.64) | 1.05 (4.82) | -1.4 | <0.001 | 0.295 |
| Unknown | 265 | 98 | | | |
| DPoAm raw residual | -0.01 (0.09) | 0.02 (0.10) | -0.03 | <0.001 | 0.324 |
| DPoAm weighted residual | 0.00 (0.09) | 0.03 (0.10) | -0.03 | <0.001 | 0.334 |
| DPoAm residual with ancestry-informative PCs | -0.01 (0.09) | 0.02 (0.09) | -0.03 | <0.001 | 0.336 |
| Unknown | 265 | 98 | | | |

Mean DNAm aging is shown in unscaled units.
[1]Mean (SD)
[2]Welch Two Sample t-test
[3]Cohen's D

**S2 Table. GrimAge aging: Multivariable regression models excluding individuals who moved 2010-2016.**

| GrimAge[1] | Total disparity[1] | Individual SES[1] | SDI[1] | Social Disorder[1] | Physical Disorder[1] | PM2.5[1] | Ozone[1] | $NO_2$[1] |
|---|---|---|---|---|---|---|---|---|
| **Race** | | | | | | | | |
| White | — | — | — | — | — | — | — | — |
| Black | 0.24** (0.10,0.37) | 0.07 (-0.06,0.19) | 0.02 (-0.11,0.15) | 0.06 (-0.07,0.18) | 0.06 (-0.07,0.19) | 0.06 (-0.07,0.18) | 0.06 (-0.07,0.19) | 0.06 (-0.07,0.18) |
| **Gender** | | | | | | | | |
| Male | | — | — | — | — | — | — | — |
| Female | | -0.70*** (-0.77,-0.62) | -0.69*** (-0.77,-0.62) | -0.69*** (-0.76,-0.62) | -0.70*** (-0.77,-0.62) | -0.70*** (-0.77,-0.62) | -0.70*** (-0.77,-0.62) | -0.70*** (-0.77,-0.62) |
| **Education** | | | | | | | | |
| College + | | — | — | — | — | — | — | — |
| Some College | | 0.16** (0.06,0.25) | 0.15* (0.06,0.25) | 0.15** (0.06,0.25) | 0.16** (0.06,0.25) | 0.15** (0.06,0.25) | 0.16** (0.06,0.25) | 0.16** (0.06,0.25) |
| High School | | 0.22*** (0.12,0.31) | 0.21*** (0.12,0.31) | 0.22*** (0.12,0.31) | 0.22*** (0.12,0.31) | 0.21*** (0.12,0.31) | 0.22*** (0.12,0.31) | 0.22*** (0.12,0.31) |
| < High School | | 0.42*** (0.28,0.56) | 0.42*** (0.28,0.56) | 0.42*** (0.28,0.56) | 0.42*** (0.28,0.56) | 0.42*** (0.28,0.56) | 0.42*** (0.28,0.56) | 0.43*** (0.28,0.57) |
| **Quartile Wealth/Income** | | | | | | | | |
| 4 | | — | — | — | — | — | — | — |
| 3 | | 0.19*** (0.10,0.29) | 0.18** (0.09,0.28) | 0.19*** (0.09,0.28) | 0.19*** (0.10,0.29) | 0.19*** (0.10,0.29) | 0.19*** (0.10,0.29) | 0.20*** (0.10,0.29) |
| 2 | | 0.44*** (0.33,0.54) | 0.41*** (0.30,0.52) | 0.43*** (0.33,0.54) | 0.44*** (0.33,0.54) | 0.44*** (0.34,0.54) | 0.44*** (0.33,0.54) | 0.44*** (0.34,0.55) |
| 1 | | 0.54*** (0.42,0.66) | 0.50*** (0.38,0.63) | 0.53*** (0.41,0.65) | 0.53*** (0.41,0.66) | 0.54*** (0.42,0.66) | 0.54*** (0.42,0.66) | 0.54*** (0.42,0.66) |
| **Neighborhood Exposure** | | | 0.05 (0.01,0.09) | 0.02 (-0.02,0.06) | 0.01 (-0.03,0.05) | 0.01 (-0.01,0.03) | 0.00 (-0.01,0.01) | 0.00 (0.00,0.01) |
| **(Intercept)** | -0.06* (-0.11,-0.02) | -0.07 (-0.15,0.01) | -0.03 (-0.12,0.05) | -0.07 (-0.15,0.02) | -0.07 (-0.15,0.02) | -0.17 (-0.36,0.02) | 0.07 (-0.29,0.43) | -0.11 (-0.22,0.01) |
| $R^2$ | 0.005 | 0.198 | 0.200 | 0.198 | 0.198 | 0.199 | 0.198 | 0.198 |
| AIC | 6,842 | 6,362 | 6,359 | 6,363 | 6,363 | 6,362 | 6,363 | 6,363 |
| No. Obs. | 2,292 | 2,292 | 2,292 | 2,292 | 2,292 | 2,292 | 2,292 | 2,292 |

Results of linear regression models with GrimAge aging as the outcome excluding 667 participants whose residential census tract changed.

[1]ß (95% confidence interval) *p<0.05; **p<0.01; ***p<0.001

**S3 Table. DPoAm aging: Multivariable regression models excluding individuals who moved 2010-2016.**

| DPoAm[1] | Total disparity[1] | Individual SES[1] | SDI[1] | Social Disorder[1] | Physical Disorder[1] | PM2.5[1] | Ozone[1] | $NO_2$[1] |
|---|---|---|---|---|---|---|---|---|
| **Race** | | | | | | | | |
| White | — | — | — | — | — | — | — | — |
| Black | 0.25** (0.12,0.39) | 0.12 (-0.02,0.26) | 0.09 (-0.06,0.23) | 0.10 (-0.04,0.24) | 0.11 (-0.03,0.25) | 0.11 (-0.03,0.25) | 0.11 (-0.03,0.25) | 0.10 (-0.04,0.25) |
| **Gender** | | | | | | | | |
| Male | | — | — | — | — | — | — | — |
| Female | | -0.17*** (-0.25,-0.09) | -0.17*** (-0.25,-0.09) | -0.16*** (-0.24,-0.08) | -0.17*** (-0.25,-0.09) | -0.17*** (-0.25,-0.09) | -0.17*** (-0.25,-0.09) | -0.17*** (-0.25,-0.09) |
| **Education** | | | | | | | | |
| College + | | — | — | — | — | — | — | — |
| Some College | | 0.15* (0.04,0.25) | 0.14* (0.04,0.25) | 0.14* (0.04,0.25) | 0.14* (0.04,0.25) | 0.14* (0.04,0.25) | 0.15* (0.04,0.25) | 0.15* (0.04,0.25) |
| High School | | 0.16* (0.05,0.26) | 0.16* (0.05,0.26) | 0.16* (0.06,0.27) | 0.16* (0.05,0.26) | 0.16* (0.05,0.26) | 0.16* (0.05,0.26) | 0.16* (0.06,0.27) |
| < High School | | 0.31*** (0.16,0.47) | 0.31*** (0.16,0.47) | 0.31*** (0.16,0.47) | 0.31*** (0.16,0.47) | 0.31*** (0.16,0.47) | 0.31*** (0.16,0.47) | 0.32*** (0.17,0.47) |
| **Quartile Wealth/Income** | | | | | | | | |
| 4 | | — | — | — | — | — | — | — |
| 3 | | 0.08 (-0.02,0.19) | 0.08 (-0.03,0.18) | 0.08 (-0.02,0.18) | 0.08 (-0.02,0.19) | 0.08 (-0.02,0.19) | 0.08 (-0.02,0.19) | 0.09 (-0.02,0.19) |
| 2 | | 0.22*** (0.11,0.33) | 0.20** (0.08,0.32) | 0.21** (0.10,0.33) | 0.22** (0.10,0.33) | 0.22*** (0.11,0.34) | 0.22** (0.11,0.33) | 0.23*** (0.11,0.34) |
| 1 | | 0.34*** (0.21,0.47) | 0.31*** (0.17,0.45) | 0.32*** (0.19,0.46) | 0.33*** (0.20,0.47) | 0.34*** (0.21,0.47) | 0.34*** (0.21,0.47) | 0.34*** (0.21,0.48) |
| **Neighborhood Exposure** | | | 0.03 (-0.01,0.08) | 0.03 (-0.01,0.07) | 0.01 (-0.03,0.06) | 0.01 (-0.01,0.03) | -0.01 (-0.02,0.00) | 0.01 (0.00,0.02) |
| **(Intercept)** | -0.06* (-0.10,-0.02) | -0.21*** (-0.30,-0.12) | -0.18*** (-0.28,-0.09) | -0.20*** (-0.29,-0.11) | -0.20*** (-0.29,-0.11) | -0.29* (-0.50,-0.08) | 0.09 (-0.31,0.49) | -0.26*** (-0.38,-0.13) |
| $R^2$ | 0.006 | 0.044 | 0.044 | 0.045 | 0.044 | 0.044 | 0.045 | 0.044 |
| AIC | 6,868 | 6,793 | 6,793 | 6,793 | 6,795 | 6,794 | 6,793 | 6,794 |
| No. Obs. | 2,292 | 2,292 | 2,292 | 2,292 | 2,292 | 2,292 | 2,292 | 2,292 |

Results of linear regression models with GrimAge aging as the outcome excluding 667 participants whose residential census tract changed.

[1]ß (95% confidence interval) *p<0.05; **p<0.01; ***p<0.001

**S4 Table: DPoAm aging: Interactions between neighborhood exposures and race**

| DPoAm[1] | SDI[1] | Social Disorder[1] | Physical Disorder[1] | PM2.5 (2014)[1] | Ozone (2014)[1] | $NO_2$ (2010)[1] | PM2.5 (2010)[1] |
|---|---|---|---|---|---|---|---|
| **Race** | | | | | | | |
| White | — | — | — | — | — | — | — |
| Black | 0.15<br>(0.00,0.31) | 0.17<br>(0.04,0.30) | 0.16<br>(0.03,0.29) | -0.81<br>(-1.5,-0.09) | -0.56<br>(-1.9,0.74) | -0.08<br>(-0.35,0.19) | -0.02<br>(-0.78,0.73) |
| **Gender** | | | | | | | |
| Male | — | — | — | — | — | — | — |
| Female | -0.20***<br>(-0.27,-0.13) | -0.20***<br>(-0.27,-0.12) | -0.20***<br>(-0.27,-0.13) | -0.20***<br>(-0.27,-0.13) | -0.20***<br>(-0.27,-0.13) | -0.20***<br>(-0.27,-0.13) | -0.20***<br>(-0.27,-0.13) |
| **Education** | | | | | | | |
| College + | — | — | — | — | — | — | — |
| Some College | 0.17**<br>(0.08,0.27) | 0.18**<br>(0.08,0.27) | 0.18**<br>(0.08,0.27) | 0.17**<br>(0.08,0.27) | 0.18***<br>(0.09,0.28) | 0.18***<br>(0.09,0.27) | 0.18**<br>(0.08,0.27) |
| High School | 0.22***<br>(0.13,0.32) | 0.23***<br>(0.13,0.32) | 0.22***<br>(0.13,0.32) | 0.23***<br>(0.13,0.32) | 0.23***<br>(0.13,0.32) | 0.23***<br>(0.13,0.32) | 0.22***<br>(0.13,0.32) |
| < High School | 0.39***<br>(0.25,0.52) | 0.39***<br>(0.25,0.53) | 0.39***<br>(0.25,0.53) | 0.39***<br>(0.26,0.53) | 0.40***<br>(0.26,0.53) | 0.39***<br>(0.26,0.53) | 0.39***<br>(0.26,0.53) |
| **Quartile Wealth/Income** | | | | | | | |
| 4 | — | — | — | — | — | — | — |
| 3 | 0.05<br>(-0.04,0.15) | 0.06<br>(-0.04,0.15) | 0.06<br>(-0.03,0.16) | 0.06<br>(-0.03,0.16) | 0.06<br>(-0.04,0.15) | 0.06<br>(-0.04,0.15) | 0.06<br>(-0.04,0.16) |
| 2 | 0.18**<br>(0.07,0.28) | 0.19**<br>(0.09,0.29) | 0.20**<br>(0.09,0.30) | 0.20***<br>(0.09,0.30) | 0.19**<br>(0.09,0.30) | 0.20**<br>(0.09,0.30) | 0.19**<br>(0.09,0.30) |
| 1 | 0.29***<br>(0.18,0.41) | 0.31***<br>(0.19,0.42) | 0.32***<br>(0.20,0.43) | 0.32***<br>(0.20,0.43) | 0.32***<br>(0.21,0.44) | 0.32***<br>(0.21,0.43) | 0.32***<br>(0.21,0.44) |
| **Neighborhood Exposure** | 0.04<br>(-0.01,0.08) | 0.02<br>(-0.02,0.06) | 0.00<br>(-0.04,0.04) | 0.00<br>(-0.02,0.01) | 0.00<br>(-0.01,0.00) | 0.00<br>(-0.01,0.01) | 0.00<br>(-0.02,0.02) |
| **Race * Neighborhood Exposure** | | | | | | | |
| Black * Neighborhood Exposure | 0.04<br>(-0.10,0.17) | 0.07<br>(-0.05,0.19) | 0.11<br>(-0.01,0.23) | 0.10*<br>(0.03,0.17) | 0.02<br>(-0.01,0.05) | 0.03<br>(0.00,0.05) | 0.02<br>(-0.05,0.09) |
| **(Intercept)** | -0.20***<br>(-0.28,-0.11) | -0.22***<br>(-0.30,-0.14) | -0.22***<br>(-0.31,-0.14) | -0.19<br>(-0.38,0.00) | -0.07<br>(-0.42,0.28) | -0.22**<br>(-0.34,-0.10) | -0.26<br>(-0.47,-0.06) |
| $R^2$ | 0.060 | 0.060 | 0.060 | 0.061 | 0.059 | 0.061 | 0.059 |
| AIC | 8,843 | 8,843 | 8,843 | 8,839 | 8,845 | 8,841 | 8,807 |

Results of linear regression models with DPoAm aging as the outcome.

[1]ß (95% confidence interval) *p<0.05; **p<0.01; ***p<0.001

**S5 Table: GrimAge aging: Interactions between neighborhood exposures and race**

| GrimAge[1] | SDI[1] | Social Disorder[1] | Physical Disorder[1] | PM2.5 (2014)[1] | Ozone (2014)[1] | $NO_2$ (2010)[1] | PM2.5 (2010)[1] |
|---|---|---|---|---|---|---|---|
| **Race** | | | | | | | |
| White | — | — | — | — | — | — | — |
| Black | 0.15 | 0.10 | 0.13 | 0.06 | -0.81 | 0.20 | -0.02 |
| | (0.01,0.29) | (-0.02,0.22) | (0.01,0.25) | (-0.61,0.73) | (-2.0,0.39) | (-0.05,0.45) | (-0.78,0.73) |
| **Gender** | | | | | | | |
| Male | — | — | — | — | — | — | — |
| Female | -0.73*** | -0.72*** | -0.73*** | -0.73*** | -0.73*** | -0.73*** | -0.20*** |
| | (-0.79,-0.66) | (-0.79,-0.66) | (-0.79,-0.66) | (-0.80,-0.67) | (-0.80,-0.67) | (-0.80,-0.67) | (-0.27,-0.13) |
| **Education** | | | | | | | |
| College + | — | — | — | — | — | — | — |
| Some College | 0.25*** | 0.25*** | 0.25*** | 0.26*** | 0.26*** | 0.26*** | 0.18** |
| | (0.17,0.34) | (0.17,0.34) | (0.17,0.34) | (0.17,0.34) | (0.17,0.35) | (0.17,0.34) | (0.08,0.27) |
| High School | 0.29*** | 0.30*** | 0.29*** | 0.29*** | 0.30*** | 0.30*** | 0.22*** |
| | (0.21,0.38) | (0.21,0.38) | (0.21,0.38) | (0.21,0.38) | (0.21,0.38) | (0.21,0.38) | (0.13,0.32) |
| < High School | 0.50*** | 0.50*** | 0.50*** | 0.51*** | 0.51*** | 0.51*** | 0.39*** |
| | (0.38,0.63) | (0.38,0.63) | (0.38,0.63) | (0.38,0.63) | (0.38,0.64) | (0.38,0.63) | (0.26,0.53) |
| **Quartile Wealth/Income** | | | | | | | |
| 4 | — | — | — | — | — | — | — |
| 3 | 0.14* | 0.15** | 0.15** | 0.15** | 0.15** | 0.15** | 0.06 |
| | (0.05,0.22) | (0.06,0.23) | (0.06,0.24) | (0.06,0.24) | (0.06,0.24) | (0.06,0.24) | (-0.04,0.16) |
| 2 | 0.37*** | 0.40*** | 0.40*** | 0.41*** | 0.41*** | 0.41*** | 0.19** |
| | (0.27,0.47) | (0.30,0.49) | (0.31,0.50) | (0.31,0.50) | (0.31,0.50) | (0.31,0.50) | (0.09,0.30) |
| 1 | 0.49*** | 0.52*** | 0.52*** | 0.53*** | 0.53*** | 0.54*** | 0.32*** |
| | (0.38,0.60) | (0.41,0.62) | (0.42,0.63) | (0.43,0.64) | (0.43,0.64) | (0.43,0.64) | (0.21,0.44) |
| **Neighborhood Exposure** | 0.07** | 0.04 | 0.02 | 0.00 | 0.00 | 0.00 | 0.00 |
| | (0.03,0.11) | (0.00,0.07) | (-0.02,0.06) | (-0.02,0.02) | (-0.01,0.01) | (-0.01,0.01) | (-0.02,0.02) |
| **Race * Neighborhood Exposure** | | | | | | | |
| Black * Neighborhood Exposure | -0.11 | 0.03 | -0.02 | 0.01 | 0.03 | -0.01 | 0.02 |
| | (-0.24,0.01) | (-0.08,0.14) | (-0.13,0.09) | (-0.06,0.07) | (-0.01,0.06) | (-0.03,0.01) | (-0.05,0.09) |
| **(Intercept)** | -0.05 | -0.08 | -0.08 | -0.11 | -0.08 | -0.11 | -0.26 |
| | (-0.13,0.03) | (-0.16,-0.01) | (-0.16,-0.01) | (-0.29,0.06) | (-0.41,0.25) | (-0.22,0.00) | (-0.47,-0.06) |
| $R^2$ | 0.216 | 0.214 | 0.213 | 0.213 | 0.214 | 0.213 | 0.059 |
| AIC | 8,385 | 8,392 | 8,395 | 8,397 | 8,394 | 8,396 | 8,807 |

Results of linear regression models with GrimAge aging as the outcome.

[1]ß (95% confidence interval) *p<0.05; **p<0.01; ***p<0.001

**S6 Table. DPoAm aging: Interactions between neighborhood and social determinants and PM2.5 pollution exposure.**

| DPoAm[1] | Race[1] | Gender[1] | Individual SES[1] | Education[1] | SDI[1] | Social Disorder[1] | Physical Disorder[1] |
|---|---|---|---|---|---|---|---|
| **Race** | | | | | | | |
| White | — | — | — | — | — | — | — |
| Black | -0.81 | 0.21** | 0.19** | 0.20** | 0.16 | 0.19* | 0.20** |
| | (-1.5,-0.09) | (0.09,0.33) | (0.07,0.32) | (0.08,0.32) | (0.03,0.29) | (0.07,0.31) | (0.08,0.32) |
| **PM2.5** | 0.00 | 0.03 | -0.03 | -0.01 | 0.01 | 0.00 | 0.00 |
| | (-0.02,0.01) | (0.01,0.06) | (-0.06,0.00) | (-0.04,0.02) | (-0.01,0.03) | (-0.01,0.02) | (-0.01,0.02) |
| **Gender** | | | | | | | |
| Male | — | — | — | — | — | — | — |
| Female | -0.20*** | 0.30 | -0.20*** | -0.20*** | -0.20*** | -0.19*** | -0.20*** |
| | (-0.27,-0.13) | (-0.05,0.65) | (-0.27,-0.13) | (-0.27,-0.13) | (-0.27,-0.13) | (-0.26,-0.12) | (-0.27,-0.13) |
| **Education** | | | | | | | |
| College + | — | — | — | — | — | — | — |
| Some College | 0.17** | 0.18*** | 0.18** | 0.02 | 0.17** | 0.18** | 0.18** |
| | (0.08,0.27) | (0.09,0.27) | (0.08,0.27) | (-0.41,0.45) | (0.08,0.27) | (0.09,0.27) | (0.08,0.27) |
| High School | 0.23*** | 0.23*** | 0.22*** | 0.20 | 0.22*** | 0.23*** | 0.23*** |
| | (0.13,0.32) | (0.13,0.32) | (0.13,0.31) | (-0.24,0.63) | (0.13,0.32) | (0.14,0.32) | (0.13,0.32) |
| < High School | 0.39*** | 0.40*** | 0.38*** | -0.33 | 0.38*** | 0.39*** | 0.39*** |
| | (0.26,0.53) | (0.26,0.53) | (0.24,0.52) | (-1.1,0.40) | (0.24,0.52) | (0.25,0.52) | (0.25,0.53) |
| **Quartile Wealth/Income** | | | | | | | |
| 4 | — | — | — | — | — | — | — |
| 3 | 0.06 | 0.06 | -0.52 | 0.06 | 0.06 | 0.06 | 0.06 |
| | (-0.03,0.16) | (-0.04,0.15) | (-1.0,-0.07) | (-0.04,0.15) | (-0.04,0.15) | (-0.04,0.15) | (-0.04,0.16) |
| 2 | 0.20*** | 0.20** | -0.07 | 0.20** | 0.18** | 0.19** | 0.19** |
| | (0.09,0.30) | (0.09,0.30) | (-0.55,0.41) | (0.09,0.30) | (0.08,0.29) | (0.09,0.29) | (0.09,0.30) |
| 1 | 0.32*** | 0.33*** | -0.30 | 0.32*** | 0.30*** | 0.31*** | 0.32*** |
| | (0.20,0.43) | (0.21,0.44) | (-0.79,0.19) | (0.20,0.43) | (0.18,0.42) | (0.19,0.42) | (0.20,0.43) |
| **Race * PM2.5** | | | | | | | |
| Black * PM2.5 | 0.10* | | | | | | |
| | (0.03,0.17) | | | | | | |
| **Gender * PM2.5** | | | | | | | |
| Female * PM2.5 | | -0.05* | | | | | |
| | | (-0.09,-0.02) | | | | | |
| **Quartile Wealth/Income * PM2.5** | | | | | | | |
| 3 * PM2.5 | | | 0.06 | | | | |
| | | | (0.01,0.11) | | | | |
| 2 * PM2.5 | | | 0.03 | | | | |
| | | | (-0.02,0.08) | | | | |
| 1 * PM2.5 | | | 0.07 | | | | |
| | | | (0.02,0.12) | | | | |
| **Education * PM2.5** | | | | | | | |
| Some College * PM2.5 | | | | 0.02 | | | |
| | | | | (-0.03,0.06) | | | |
| High School * PM2.5 | | | | 0.00 | | | |
| | | | | (-0.04,0.05) | | | |
| < High School * PM2.5 | | | | 0.08 | | | |
| | | | | (0.00,0.15) | | | |
| **Social Deprivation Index** | | | | | -0.14 | | |
| | | | | | (-0.32,0.05) | | |

| DPoAm[1] | Race[1] | Gender[1] | Individual SES[1] | Education[1] | SDI[1] | Social Disorder[1] | Physical Disorder[1] |
|---|---|---|---|---|---|---|---|
| **Social Deprivation Index * PM2.5** | | | | | 0.02 (0.00,0.04) | | |
| **Social Disorder** | | | | | | -0.10 (-0.28,0.09) | |
| **Social Disorder * PM2.5** | | | | | | 0.01 (-0.01,0.03) | |
| **Physical Disorder** | | | | | | | -0.08 (-0.27,0.11) |
| **Physical Disorder * PM2.5** | | | | | | | 0.01 (-0.01,0.03) |
| **(Intercept)** | -0.19 (-0.38,0.00) | -0.54*** (-0.81,-0.27) | 0.08 (-0.22,0.39) | -0.15 (-0.45,0.15) | -0.27* (-0.46,-0.07) | -0.26* (-0.45,-0.07) | -0.26* (-0.45,-0.07) |
| $R^2$ | 0.061 | 0.062 | 0.062 | 0.060 | 0.061 | 0.060 | 0.059 |
| AIC | 8,839 | 8,838 | 8,841 | 8,846 | 8,842 | 8,844 | 8,848 |

Results of linear regression models with DPoAm aging as the outcome.

[1]ß (95% confidence interval) *p<0.05; **p<0.01; ***p<0.001

**S7 Table: Threefold decomposition of individual and neighborhood contributions to racial disparity in DNAm aging.** Full results of decomposition for GrimAge and DPoAm.

| | | GrimAge | | | DPoAm | | |
|---|---|---|---|---|---|---|---|
| Variable | Component | Percent | Estimate | 95% CI | Percent | Estimate | 95% CI |
| Overall | Endowments | **58.82** | **0.18** | **(0.11, 0.25)** | **41.57** | **0.14** | **(0.07, 0.21)** |
| | Coefficients | **67.84** | **0.21** | **(0.05, 0.36)** | 30.53 | 0.10 | (-0.05, 0.25) |
| | Interaction | -26.66 | -0.08 | (-0.23, 0.07) | 27.90 | 0.09 | (-0.05, 0.24) |
| Intercept | Endowments | 0.00 | 0.00 | (0.00, 0.00) | 0.00 | 0.00 | (0.00, 0.00) |
| | Coefficients | -166.61 | -0.51 | (-1.81, 0.80) | **-407.10** | **-1.37** | **(-2.57, -0.17)** |
| | Interaction | -0.00 | -0.00 | (-0.00, 0.00) | -0.00 | -0.00 | (-0.00, 0.00) |
| Female | Endowments | **-27.40** | **-0.08** | **(-0.12, -0.05)** | **-6.80** | **-0.02** | **(-0.04, -0.01)** |
| | Coefficients | 30.35 | 0.09 | (-0.02, 0.21) | -13.00 | -0.04 | (-0.17, 0.08) |
| | Interaction | 5.93 | 0.02 | (-0.01, 0.04) | -2.54 | -0.01 | (-0.03, 0.02) |
| Education: Some College | Endowments | 0.23 | 0.00 | (-0.00, 0.00) | 0.38 | 0.00 | (-0.00, 0.00) |
| | Coefficients | 13.44 | 0.04 | (-0.00, 0.09) | **22.63** | **0.08** | **(0.03, 0.12)** |
| | Interaction | -0.87 | -0.00 | (-0.01, 0.01) | -1.46 | -0.00 | (-0.02, 0.01) |
| High School | Endowments | -0.01 | -0.00 | (-0.00, 0.00) | -0.03 | -0.00 | (-0.00, 0.00) |
| | Coefficients | 10.98 | 0.03 | (-0.02, 0.08) | 1.66 | 0.01 | (-0.05, 0.06) |
| | Interaction | -0.06 | -0.00 | (-0.01, 0.01) | -0.01 | -0.00 | (-0.00, 0.00) |
| Less than High School | Endowments | **11.10** | **0.03** | **(0.02, 0.05)** | **6.73** | **0.02** | **(0.01, 0.04)** |
| | Coefficients | **-7.84** | **-0.02** | **(-0.04, -0.00)** | -4.80 | -0.02 | (-0.04, 0.00) |
| | Interaction | **-10.10** | **-0.03** | **(-0.06, -0.00)** | -6.19 | -0.02 | (-0.05, 0.01) |
| Wealth/Income: 3rd Quartile | Endowments | 2.02 | 0.01 | (-0.00, 0.01) | 1.52 | 0.01 | (-0.00, 0.01) |
| | Coefficients | -13.62 | -0.04 | (-0.09, 0.01) | -11.83 | -0.04 | (-0.10, 0.02) |
| | Interaction | 6.03 | 0.02 | (-0.01, 0.04) | 5.24 | 0.02 | (-0.01, 0.04) |
| 2nd Quartile | Endowments | 1.01 | 0.00 | (-0.00, 0.01) | 0.13 | 0.00 | (-0.00, 0.00) |
| | Coefficients | 1.12 | 0.00 | (-0.04, 0.05) | 7.01 | 0.02 | (-0.02, 0.07) |
| | Interaction | 0.17 | 0.00 | (-0.01, 0.01) | 1.07 | 0.00 | (-0.00, 0.01) |
| Lowest Quartile | Endowments | **21.49** | **0.07** | **(0.04, 0.09)** | **14.39** | **0.05** | **(0.02, 0.08)** |
| | Coefficients | 7.37 | 0.02 | (-0.01, 0.06) | 5.02 | 0.02 | (-0.02, 0.05) |
| | Interaction | 12.91 | 0.04 | (-0.02, 0.10) | 8.79 | 0.03 | (-0.03, 0.09) |
| Social Deprivation Index | Endowments | **21.09** | **0.06** | **(0.02, 0.11)** | 7.02 | 0.02 | (-0.03, 0.08) |
| | Coefficients | 11.73 | 0.04 | (-0.01, 0.08) | 3.04 | 0.01 | (-0.03, 0.05) |
| | Interaction | -40.75 | -0.12 | (-0.27, 0.02) | -10.57 | -0.04 | (-0.18, 0.11) |
| Social Disorder | Endowments | 7.26 | 0.02 | (-0.01, 0.05) | 7.20 | 0.02 | (-0.01, 0.06) |
| | Coefficients | 1.22 | 0.00 | (-0.02, 0.03) | 2.18 | 0.01 | (-0.02, 0.03) |
| | Interaction | -4.37 | -0.01 | (-0.09, 0.06) | -7.81 | -0.03 | (-0.11, 0.06) |
| Physical Disorder | Endowments | -5.48 | -0.02 | (-0.05, 0.02) | -4.77 | -0.02 | (-0.06, 0.02) |
| | Coefficients | -0.52 | -0.00 | (-0.03, 0.02) | -4.35 | -0.01 | (-0.04, 0.01) |
| | Interaction | 1.92 | 0.01 | (-0.08, 0.10) | 16.02 | 0.05 | (-0.04, 0.15) |
| PM2.5 | Endowments | -0.09 | -0.00 | (-0.02, 0.02) | -2.70 | -0.01 | (-0.03, 0.01) |
| | Coefficients | -16.62 | -0.05 | (-0.62, 0.52) | 144.90 | 0.49 | (-0.10, 1.07) |
| | Interaction | -1.60 | -0.00 | (-0.06, 0.05) | 13.95 | 0.05 | (-0.01, 0.10) |
| Ozone | Endowments | -0.54 | -0.00 | (-0.01, 0.01) | 0.91 | 0.00 | (-0.01, 0.02) |
| | Coefficients | 160.63 | 0.49 | (-0.67, 1.65) | 223.56 | 0.75 | (-0.35, 1.86) |
| | Interaction | -5.46 | -0.02 | (-0.06, 0.02) | -7.61 | -0.03 | (-0.06, 0.01) |
| $NO_2$ | Endowments | 1.62 | 0.00 | (-0.02, 0.03) | 4.18 | 0.01 | (-0.01, 0.04) |
| | Coefficients | 29.26 | 0.09 | (-0.10, 0.28) | 34.05 | 0.11 | (-0.07, 0.30) |
| | Interaction | 9.76 | 0.03 | (-0.03, 0.09) | 11.36 | 0.04 | (-0.02, 0.10) |
